# Long-term Air Pollution Exposure are Associated with Higher Incidence of ST-elevation Myocardial Infarction and In-hospital Cardiogenic Shock

**DOI:** 10.1101/2023.06.08.23291174

**Authors:** Jinah Cha, Se Yeon Choi, Seung-Woon Rha, Byoung Geol Choi, Jae Kyeong Byun, Sujin Hyun, Min Woo Lee, Jaeho Kang, Wonsang Chu, Soohyung Park, Eun Jin Park, Dong Oh Kang, Cheol Ung Choi, Suhng Wook Kim, Myung ho Jeong, the other Korea Acute Myocardial Infarction Registry (KAMIR) investigators

## Abstract

**Background:** Previous studies have reported the association between myocardial infarction (MI) and air pollution (AP). However, limited information is available regarding the long-term effects of AP on the relative incidence rates of ST-elevation MI (STEMI) and Non-ST-elevation MI (NSTEMI).

**Methods:** Study subjects were enrolled from the Korea Acute MI registry (KAMIR) and KAMIR-National Institutes of Health (NIH). A total of 45,619 eligible patients with AMI were enrolled between January 2006 and December 2015. We investigated the association between long-term exposure to AP and compared the incidence of STEMI with those of NSTEMI. Mixed-effect regression models were used to examine the association between the annual average ambient AP before MI onset and the incidence of STEMI compared with that of NSTEMI, and to evaluate the association of AP with the incidence of in-hospital cardiogenic shock.

**Results:** After mixed-effect regression model analysis, each 1 μg/m^3^ increase in particulate matter (PM) 10 µm or less in diameter (PM_10_) was associated with increased incidence of STEMI compared with NSTEMI (odds ratio [OR]:1.009, 95% Confidence Interval [CI]: 1.002–1.016; p = 0.012). For in-hospital cardiogenic shock complication, each 1 μg/m^3^ increase in PM_10_ and 1 part per billion increase in SO_2_ were associated with increased risk, PM_10_ (OR:1.033, 95% CI, 1.018–1.050; p < 0.001), SO_2_ (OR:1.104, 95% CI, 1.006–1.212; p = 0.037), respectively.

**Conclusion:** A high concentration of air pollutants, particularly PM_10,_ was an environmental risk factor for an increase incidence of development of STEMI. PM_10_ and SO_2_ were found to be risk factors for in-hospital cardiogenic shock complications. Policy-level strategies and clinical efforts to reduce AP exposure are necessary to prevent the incidence of STEMI and severe cardiovascular complications.

## Clinical Perspective What is new?

– This study determined the long-term association between air pollutants and the relative incidence of STEMI and NSTEMI using a large patient registry dataset.

– Air pollution, particularly involving PM_10,_ was associated with an increased risk of STEMI compared with NSTEMI.

– In patients with Acute Myocardial Infarction (AMI), PM_10_ and SO_2_ were found to be risk factors for the development of in-hospital cardiogenic shock complications.

## What are the clinical implications?

– The findings of this study should be considered a milestone in developing a policy to reduce AP exposure and prevent STEMI incidence in high-risk MI patients.

– Clinical efforts are necessary to reduce the incidence rates of adverse cardiovascular complications such as cardiogenic shock.

– Mixed-effect regression model analysis for AP exposure is useful in providing information that can enable early identification of STEMI patients and those with severe complication cardiogenic shock risks.

## Non-standard Abbreviations and Acronyms

ACS acute coronary syndrome

AMI acute myocardial infarction

AP air pollution

BMI body mass index

CAD coronary artery disease

CI confidence intervals

CO carbon monoxide

CVA cerebrovascular disease

DL dyslipidemia

DM diabetes mellitus

HF heart failure

HTN hypertension

IHD Ischemic heart diseases

LVEF left ventricular ejection fraction MI myocardial infarction

MVD multivessel coronary artery disease NO_2_ nitric dioxide

NSTEMI Non-ST-elevation myocardial infarction O_3_ ozone

OR odds ratios

PCI percutaneous coronary intervention

PM_10_ particulate matter 10 µm or less in diameter PM_2.5_ particulate matter 2.5 µm or less in diameter ppm part per million

SO_2_ sulfur dioxide

STEMI ST-elevation myocardial infarction

## Introduction

Ischemic heart diseases (IHD) including acute myocardial infarction (AMI) are a public health burden and are a leading cause of mortality and morbidity worldwide. AMI is a major cause of mortality in the Asia-Pacific region.^1–4^ In particular, in patients with coronary artery disease (CAD), air pollution (AP) is related to complications such as increased hospitalization, re-admission, and early mortality.^5–7^ Exposure to highly polluted air is one of the environmental factors that triggers AMI.^8^ There is a relationship between short and long-term effects of exposure to AP; however, long-term exposure effects outnumber the sum of all short-term effects.^9^ Most of the studies have focused on the association between the effects of short-term exposure to AP and AMI.^8, 10, 11^ Our previous studies demonstrated that AP exposure was associated with overall adverse clinical outcomes, including mortality, in AMI patients based on short and long-term exposure and follow-up periods. ^12, 13^ However, only few studies have reported on long-term AP exposure along with comparison between the relative incidence of ST-elevation myocardial infarction(MI, STEMI) and that of Non-ST-elevation myocardial infarction (Non-STEMI) and associated adverse effects related to mortality.

This study is an extension of our previous study and aimed to clarify the association between the long-term average concentration of AP and the relative risk of developing a STEMI compared to a NSTEMI, as well as the relationship between the annual average concentration of AP and cardiogenic shock, which was observed to be high in STEMI patients.

## Methods

### Study Protocols and Population

The study subjects were enrolled in the Korea AMI registry (KAMIR) and KAMIR-National Institutes of Health (NIH). The KAMIR study protocol has been introduced previously.^14^ KAMIR and KAMIR-NIH are nationwide prospective multicenter registration study series that aim to establish treatment guidelines and derive risk factors through the analysis of various clinical characteristics and follow-up of Korean AMI patients since October 2005 onwards. A flowchart of the study is shown in Figure 1. A total of 50,130 patients with AMI were enrolled in the KAMIR and KAMIR-NIH between January 2006 and December 2015. The exclusion criteria were as follows: (1) date of symptom onset before 2006, (2) missing date of symptom onset, (3) age < 18 years, and (4) no final diagnosis of MI at discharge.

**Figure 1.**
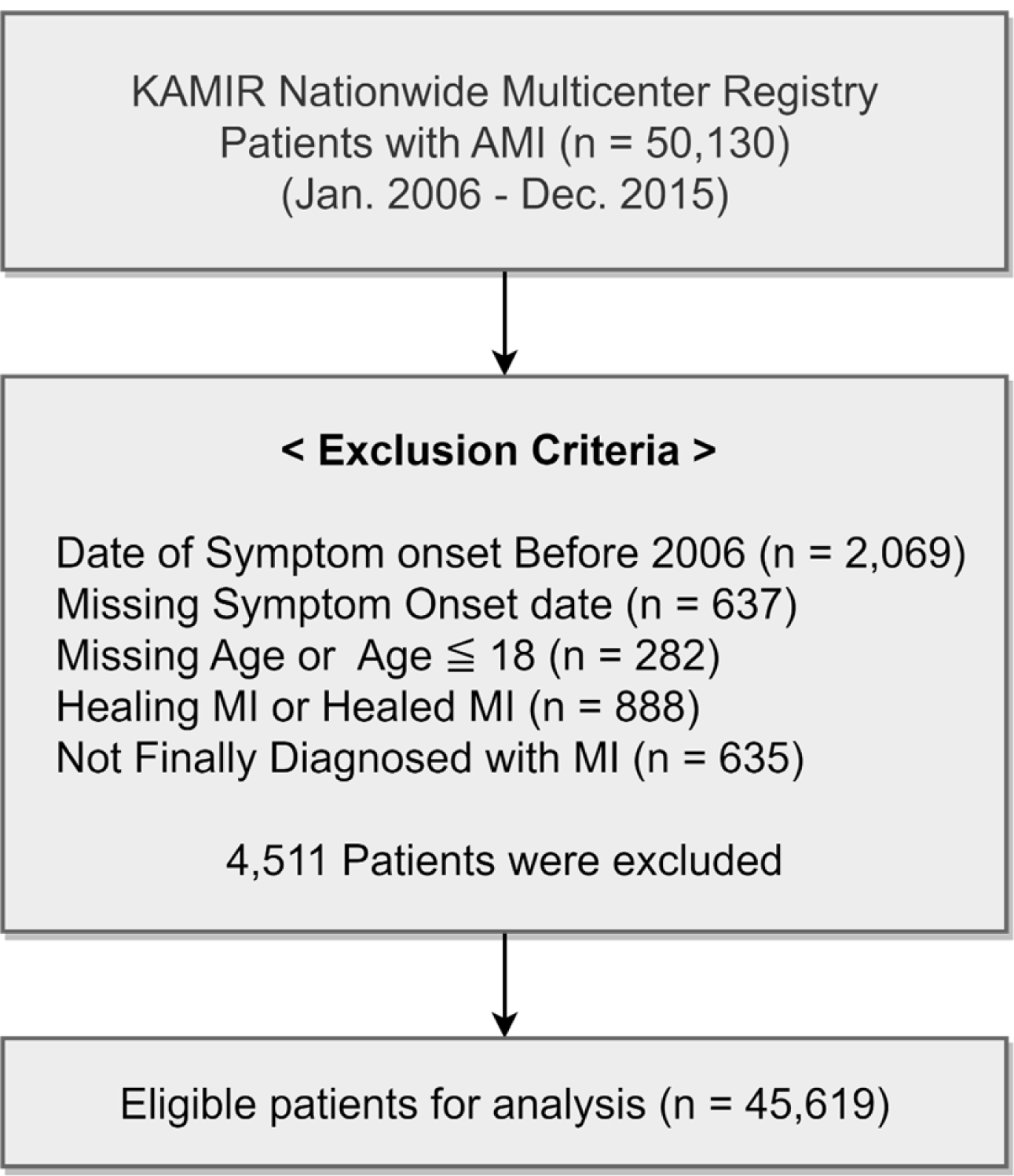
Study flow chart of patient enrollment. AMI, acute myocardial infarction; KAMIR, Korea Acute Myocardial Infarction Registry; MI, myocardial infarction

### Ethics approval

This study was approved by the Institutional Review Board (IRB) of Korea University Guro Hospital (KUGH, #2016GR0740) and was conducted in accordance with the principles of the Declaration of Helsinki.

### AP measurement

Hourly AP concentrations were provided by the Korean Ministry of Environment (http://www.airkorea.or.kr). In 2001, 329 monitoring stations nationwide began measuring the concentration of air pollutants. Measurement of air pollutants involved the β-ray absorption method for particulate matter (PM) 10 µm or less in diameter (PM_10_), the non-dispersive infrared method for carbon monoxide (CO), the pulse ultraviolet fluorescence method for sulfur dioxide (SO_2_), the chemiluminescence method for nitric dioxide (NO_2_), and the ultraviolet photometric method for ozone (O_3_). The concentration measurement of PM 2.5 µm or less in diameter (PM_2.5_) began in January 2015; therefore, annual average concentration values were not available during the patient enrollment period (2006–2015) and was excluded.

We transformed collected data into the daily average value, and then, the annual average value of air pollutants before the symptom day was calculated the way previous research was performed.^13^ Each monitoring station was matched by the closest distance in a straight line to 68 hospitals registered in KAMIR to measure individual exposure concentration of air pollutants. Monitoring stations were selected based on hospital admission addresses for the following reasons: (1) Patient addresses were not included in the multicenter registry data. (2) As AMI is an emergency, it is assumed that the patient was admitted to an emergency room close to the workplace and residence at the time of symptom onset. If a pollutant measurement was missed due to a connection error with a monitoring station, the measurement of the next-nearest monitoring station was assigned. Symptom date was defined as the first occurrence of MI-related symptoms such as chest pain or dyspnea.

### Study Definitions

The diagnosis of AMI was defined as an elevation in cardiac biomarkers (creatinine kinase-MB, and troponin I, or T) with typical changes on 12 leads electrocardiogram (ECG) or clinical symptoms. STEMI was diagnosed as a new ST-elevation segment measuring ≥1 mm from ≥2 contiguous leads on ECG. Patients with positive cardiac biomarkers but without ECG findings of STEMI were defined as NSTEMI. Cardiogenic shock was defined as a systolic blood pressure <90 mmHg for >30 min, the need for supportive management to maintain systolic blood pressure >90 mmHg, and clinical signs of pulmonary congestion.

Individual cardiovascular risk factors, including hypertension (HTN), dyslipidemia (DL), diabetes mellitus (DM), prior cardiovascular disease, heart failure (HF), prior cerebrovascular disease (CVA), and smoking history, were based on self reports by the patient.

### Statistical Analysis

All statistical analyses were performed using R version 4.1.2. (R Core Team, 2021; R: Language and Environment for Statistical Computing; R Foundation for Statistical Computing, Vienna, Austria, URL: https://www.R-project.org/).

We compared the clinical and angiographic characteristics using a X^2^ test or Fisher’s exact test for categorical variables and Student’s t-test or Mann–Whitney rank test for continuous variables. Categorical data were expressed as percentages, and continuous variables were described as mean ± standard deviation.

We used generalized logistic mixed effect models with a random effect term for hospitals to examine the associations of each air pollutant with the incidence rate of STEMI and cardiogenic shock complication rates, and to account for hospital and regional effects such as accessibility and treatment plans. Using a multivariable model, we adjusted for potential confounding factors including: age, sex, body mass index (BMI), smoking status, HTN, DM, DL, stroke, HF, previous IHD, and symptom date. STEMI status, percutaneous coronary intervention (PCI), and left ventricular ejection fraction (LVEF), were used to analyze the incidence of cardiogenic shock complications.

In the subgroup analysis, several stratified analyses were performed to test the potential effect modifications of the following factors, by including an interaction term: for the STEMI group analysis, the following terms were used: age, sex, HTN, DM, DL, CVA, HF, prior IHD, smoking, family history of CAD. In the cardiogenic shock group, STEMI status, PCI, and LVEF were added. The results were presented as adjusted odds ratios (OR) for logistic regression with corresponding 95% confidence intervals (CI). Statistical significance was defined as a p-value < 0.05.

## Results

A total of 45,619 patients with AMI were enrolled in our study. Of these, 20,526 were patients with NSTEMI and 25,093 were patients with STEMI. In our study population, compared with patients with NSTEMI, patients with STEMI were younger, male, had a higher smoking status, and had fewer underlying chronic diseases, such as DM, HTN, and DL. Moreover, patients with STEMI had more Killip class IV and a low LVEF and those were less likely to have a history of cardiovascular diseases, such as HF, CVA, and previous IHD. Among the angiographic parameters, the STEMI group had more PCI as the initial treatment for MI, lower multivessel coronary artery disease (MVD), and the left main artery as the culprit lesion. The cardiogenic shock complication rate during the index hospitalization was significantly higher in the STEMI group (Table 1). Therefore, we analyzed the link between cardiogenic shock events, which are severe cardiovascular complications, and annual AP exposure (Table 4).

**Table 1.**
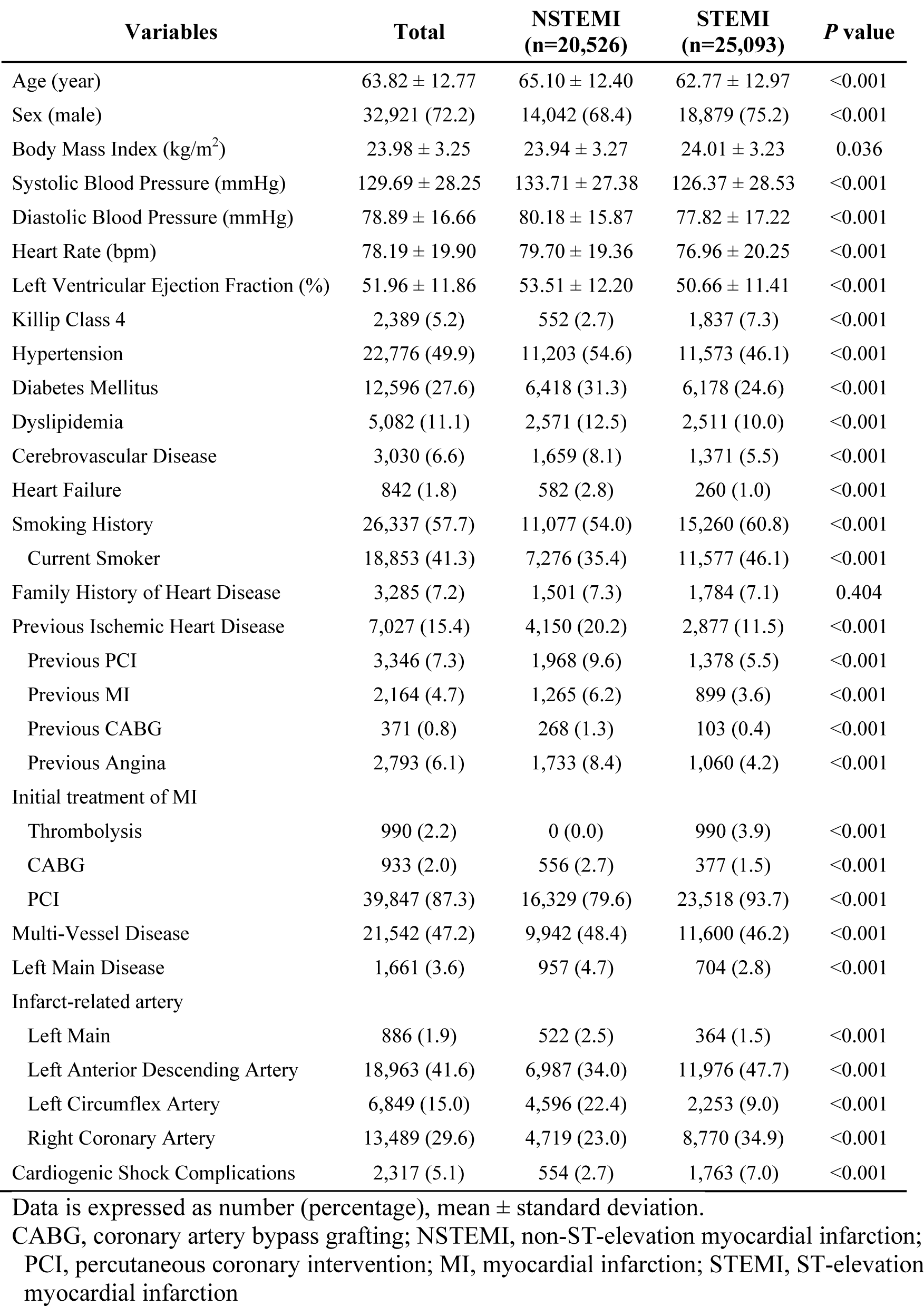
Baseline characteristics

| Variables | Total | NSTEMI<br>(n=20,526) | STEMI<br>(n=25,093) | P value |
| --- | --- | --- | --- | --- |
| Age (year) | 63.82 ± 12.77 | 65.10 ± 12.40 | 62.77 ± 12.97 | <0.001 |
| Sex (male) | 32,921 (72.2) | 14,042 (68.4) | 18,879 (75.2) | <0.001 |
| Body Mass Index (kg/m<sup>2</sup>) | 23.98 ± 3.25 | 23.94 ± 3.27 | 24.01 ± 3.23 | 0.036 |
| Systolic Blood Pressure (mmHg) | 129.69 ± 28.25 | 133.71 ± 27.38 | 126.37 ± 28.53 | <0.001 |
| Diastolic Blood Pressure (mmHg) | 78.89 ± 16.66 | 80.18 ± 15.87 | 77.82 ± 17.22 | <0.001 |
| Heart Rate (bpm) | 78.19 ± 19.90 | 79.70 ± 19.36 | 76.96 ± 20.25 | <0.001 |
| Left Ventricular Ejection Fraction (%) | 51.96 ± 11.86 | 53.51 ± 12.20 | 50.66 ± 11.41 | <0.001 |
| Killip Class 4 | 2,389 (5.2) | 552 (2.7) | 1,837 (7.3) | <0.001 |
| Hypertension | 22,776 (49.9) | 11,203 (54.6) | 11,573 (46.1) | <0.001 |
| Diabetes Mellitus | 12,596 (27.6) | 6,418 (31.3) | 6,178 (24.6) | <0.001 |
| Dyslipidemia | 5,082 (11.1) | 2,571 (12.5) | 2,511 (10.0) | <0.001 |
| Cerebrovascular Disease | 3,030 (6.6) | 1,659 (8.1) | 1,371 (5.5) | <0.001 |
| Heart Failure | 842 (1.8) | 582 (2.8) | 260 (1.0) | <0.001 |
| Smoking History | 26,337 (57.7) | 11,077 (54.0) | 15,260 (60.8) | <0.001 |
| Current Smoker | 18,853 (41.3) | 7,276 (35.4) | 11,577 (46.1) | <0.001 |
| Family History of Heart Disease | 3,285 (7.2) | 1,501 (7.3) | 1,784 (7.1) | 0.404 |
| Previous Ischemic Heart Disease | 7,027 (15.4) | 4,150 (20.2) | 2,877 (11.5) | <0.001 |
| Previous PCI | 3,346 (7.3) | 1,968 (9.6) | 1,378 (5.5) | <0.001 |
| Previous MI | 2,164 (4.7) | 1,265 (6.2) | 899 (3.6) | <0.001 |
| Previous CABG | 371 (0.8) | 268 (1.3) | 103 (0.4) | <0.001 |
| Previous Angina | 2,793 (6.1) | 1,733 (8.4) | 1,060 (4.2) | <0.001 |
| Initial treatment of MI |  |  |  |  |
| Thrombolysis | 990 (2.2) | 0 (0.0) | 990 (3.9) | <0.001 |
| CABG | 933 (2.0) | 556 (2.7) | 377 (1.5) | <0.001 |
| PCI | 39,847 (87.3) | 16,329 (79.6) | 23,518 (93.7) | <0.001 |
| Multi-Vessel Disease | 21,542 (47.2) | 9,942 (48.4) | 11,600 (46.2) | <0.001 |
| Left Main Disease | 1,661 (3.6) | 957 (4.7) | 704 (2.8) | <0.001 |
| Infarct-related artery |  |  |  |  |
| Left Main | 886 (1.9) | 522 (2.5) | 364 (1.5) | <0.001 |
| Left Anterior Descending Artery | 18,963 (41.6) | 6,987 (34.0) | 11,976 (47.7) | <0.001 |
| Left Circumflex Artery | 6,849 (15.0) | 4,596 (22.4) | 2,253 (9.0) | <0.001 |
| Right Coronary Artery | 13,489 (29.6) | 4,719 (23.0) | 8,770 (34.9) | <0.001 |
| Cardiogenic Shock Complications | 2,317 (5.1) | 554 (2.7) | 1,763 (7.0) | <0.001 |

Data is expressed as number (percentage), mean ± standard deviation.
CABG, coronary artery bypass grafting; NSTEMI, non-ST-elevation myocardial infarction;
PCI, percutaneous coronary intervention; MI, myocardial infarction; STEMI, ST-elevation
myocardial infarction

In Table 2, we observed the median value of annual average concentrations was 0.049 part per million (ppm) for SO_2_, 0.6088 ppm for CO, 0.0211 ppm for O_3_, 0.0259 ppm for NO_2_, and 50.53 μg/m^3^ for PM_10_.

**Table 2.** Distribution for annual average air pollution concentration before symptom date.

|  | SO<sub>2</sub> (ppm) | CO (ppm) | O<sub>3</sub> (ppm) | NO<sub>2</sub> (ppm) | PM<sub>10</sub> (μg/m<sup>3</sup>) |
| --- | --- | --- | --- | --- | --- |
| Min. | 0.0014 | 0.2028 | 0.0037 | 0.0070 | 18.25 |
| Q1 | 0.0041 | 0.5312 | 0.0176 | 0.0218 | 44.65 |
| Median | 0.0049 | 0.6088 | 0.0211 | 0.0259 | 50.53 |
| Q3 | 0.0061 | 0.7211 | 0.0244 | 0.0342 | 58.67 |
| Max. | 0.0136 | 1.4976 | 0.0437 | 0.0812 | 99.14 |
| IQR | 0.0020 | 0.1899 | 0.0068 | 0.0124 | 14.02 |
| Mean | 0.0053 | 0.6276 | 0.0210 | 0.0284 | 52.57 |

CO, carbon monoxide; IQR, interquartile range; NO<sub>2</sub>, nitric dioxide; O<sub>3</sub>, ozone; PM<sub>10</sub>, particulate matter 10 μm or less in diameter; ppm, part per million; Q1, 1<sup>st</sup> quintile; Q3, 3<sup>rd</sup> quintile. SO<sub>2</sub>, sulfur dioxide.

In the Spearman rank correlation analysis using the average annual concentrations after the symptom date, most air pollutants showed a positive correlation (r = 0.178–0.467); however, O_3,_ and other air pollutants, showed a negative correlation (r = −0.265–-0.609; Figure 2).

**Figure 2.**
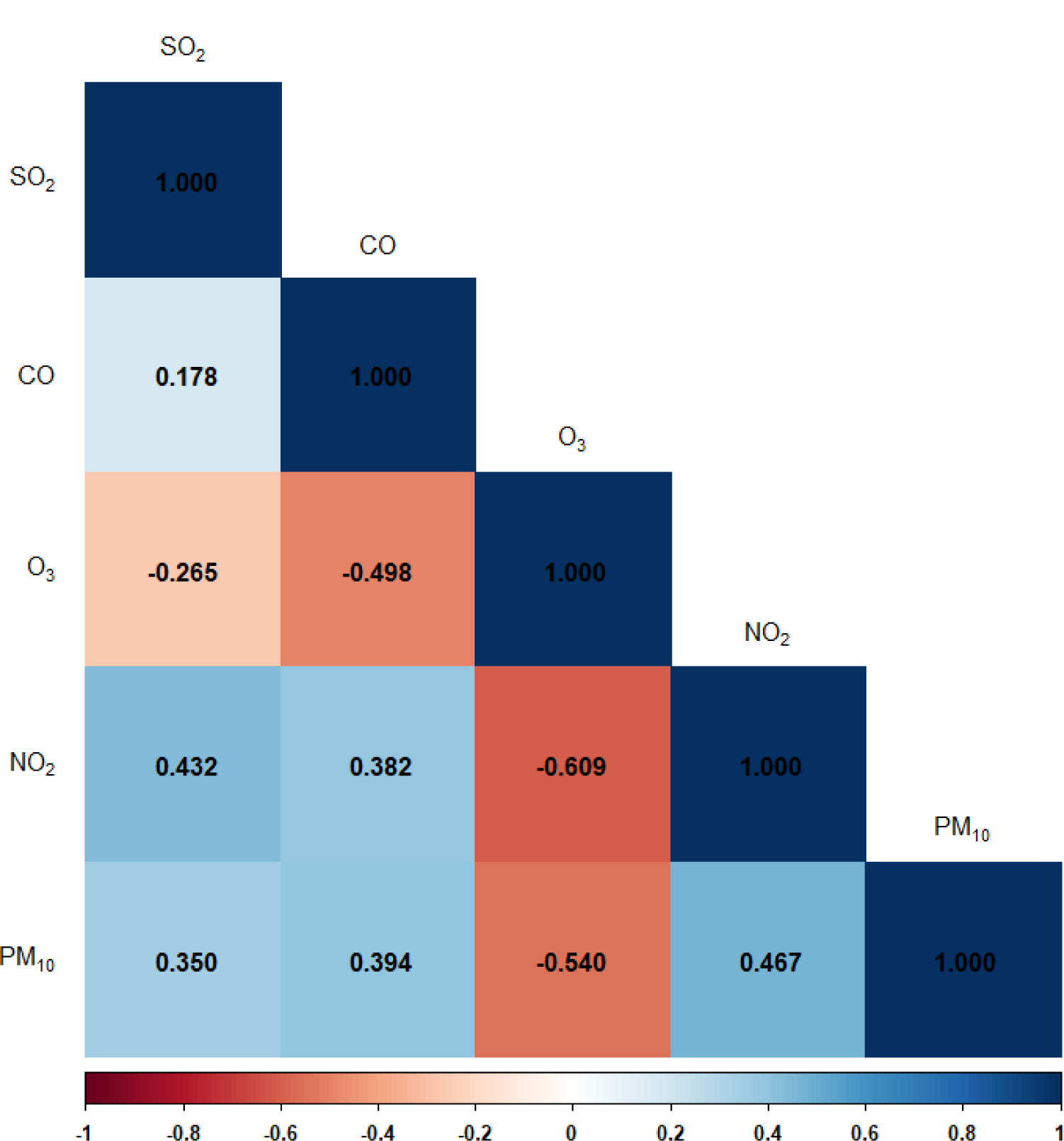
Spearman correlation coefficients for annual average concentrations of air pollutants. CO, carbon monoxide; NO_2_, nitric dioxide; PM_10_, particulate matter 10 µm or less in diameter.

After mixed-effect regression model analysis, no difference was observed for most air pollutants except PM_10_, which was associated with increased incidence of STEMI compared with NSTEMI for each 1 μg/m^3^ increase (odds ratio [OR]:1.009, 95% Confidence Interval [CI]: 1.002–1.016; p=0.012; Table 3). For in-hospital cardiogenic shock complication, each 1 μg/m^3^ increase of PM_10_ and each 1 part per billion (ppb) increase of SO_2_ were associated with increased risk: PM_10_ (OR:1.033, 95% CI: 1.018–1.050; p<0.001), SO_2_ (OR:1.104, 95% CI: 1.006–1.212; p=0.037), respectively. In contrast, for each 1 ppb increase in O_3_ was negatively correlated with cardiogenic shock (OR, 0.891; 95% CI:0.857– 0.928; p<0.001; Table 4).

**Table 3.** Univariate and multivariate regression analysis of the incidence of STEMI compared with NSTEMI regarding annual average concentration of each air pollutant before symptom date.

|  | Univariate |  | Multivariate |  |
| --- | --- | --- | --- | --- |
|  | OR (95% CI) | *P* value | OR (95% CI) | *P* value |
| SO<sub>2</sub> (ppb) | 1.044 (0.998 - 1.093) | 0.060 | 1.041 (0.995 - 1.089) | 0.084 |
| CO (ppm) | 1.037 (0.978 - 1.100) | 0.225 | 1.036 (0.977 - 1.099) | 0.236 |
| O<sub>3</sub> (ppb) | 0.990 (0.972 - 1.009) | 0.293 | 0.989 (0.971 - 1.007) | 0.239 |
| NO<sub>2</sub> (ppb) | 1.000 (0.987 - 1.012) | 0.943 | 0.999 (0.987 - 1.012) | 0.914 |
| PM<sub>10</sub> (μg/m<sup>3</sup>) | 1.008 (1.001 - 1.014) | 0.027 | 1.009 (1.002 - 1.016) | 0.012 |

Adjusted for Age, body mass index, diabetes mellitus, dyslipidemia, heart failure, hypertension, previous ischemic heart disease, sex, smoking status, stroke, symptom onset. CI, confidence interval; CO, carbon monoxide; NO<sub>2</sub>, nitric dioxide; O<sub>3</sub>, ozone; OR, odds ratio; PM<sub>10</sub>, particulate matter 10 μm or less in diameters; ppb, part per billion; ppm, part per million; SO<sub>2</sub>, sulfur dioxide.

**Table 4.**
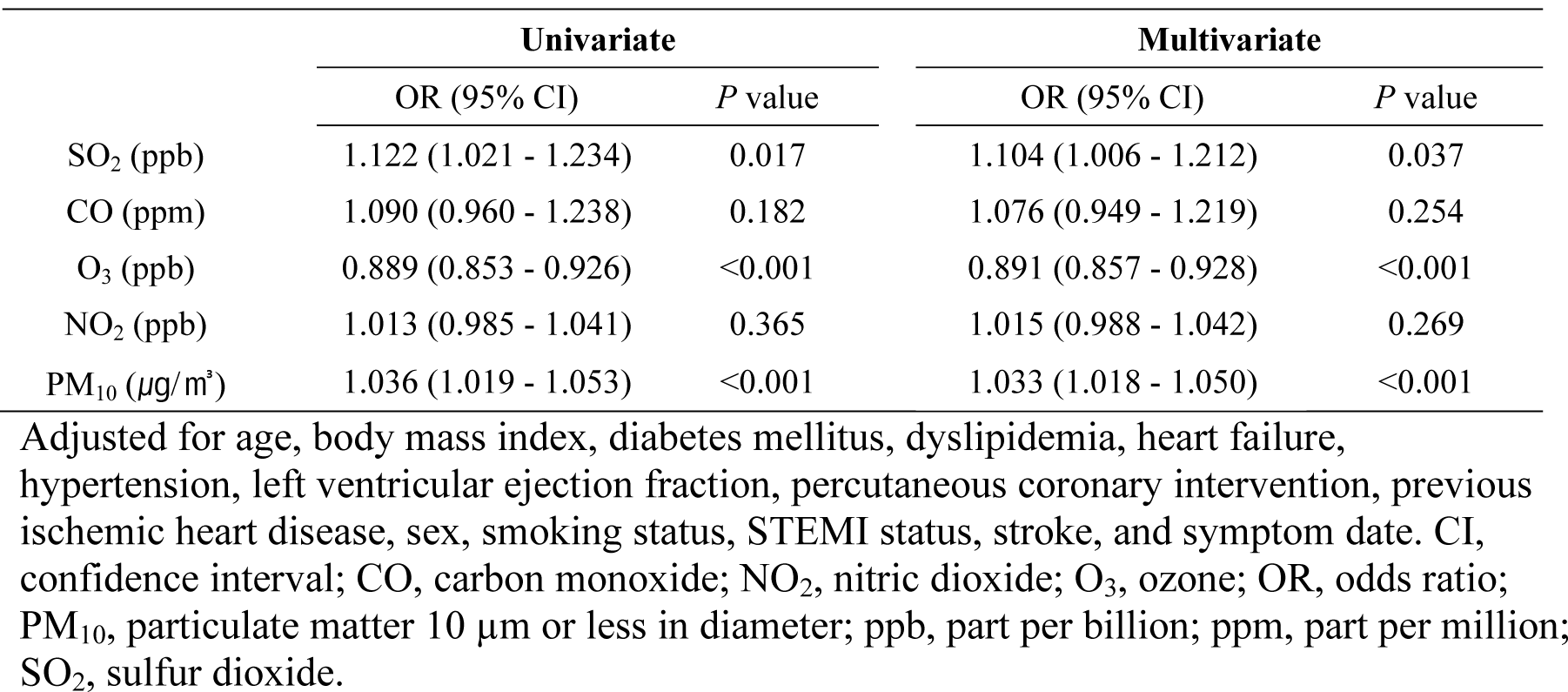
Univariate and multivariate regression analysis between the incidence of cardiogenic shock events and the annual average concentration of each air pollutant before the symptom date of myocardial infarction.

|  | Univariate |  | Multivariate |  |
| --- | --- | --- | --- | --- |
|  | OR (95% CI) | *P* value | OR (95% CI) | *P* value |
| SO<sub>2</sub> (ppb) | 1.122 (1.021 - 1.234) | 0.017 | 1.104 (1.006 - 1.212) | 0.037 |
| CO (ppm) | 1.090 (0.960 - 1.238) | 0.182 | 1.076 (0.949 - 1.219) | 0.254 |
| O<sub>3</sub> (ppb) | 0.889 (0.853 - 0.926) | <0.001 | 0.891 (0.857 - 0.928) | <0.001 |
| NO<sub>2</sub> (ppb) | 1.013 (0.985 - 1.041) | 0.365 | 1.015 (0.988 - 1.042) | 0.269 |
| PM<sub>10</sub> (μg/m<sup>3</sup>) | 1.036 (1.019 - 1.053) | <0.001 | 1.033 (1.018 - 1.050) | <0.001 |

Adjusted for age, body mass index, diabetes mellitus, dyslipidemia, heart failure, hypertension, left ventricular ejection fraction, percutaneous coronary intervention, previous ischemic heart disease, sex, smoking status, STEMI status, stroke, and symptom date. CI, confidence interval; CO, carbon monoxide; NO<sub>2</sub>, nitric dioxide; O<sub>3</sub>, ozone; OR, odds ratio; PM<sub>10</sub>, particulate matter 10 μm or less in diameter; ppb, part per billion; ppm, part per million; SO<sub>2</sub>, sulfur dioxide.

When STEMI and each air pollutant were analyzed in subgroups, the results showed there was a significant association with a decrease in STEMI incidence for every 1 ppb increase of NO_2_ in CVA patients. In the absence of HTN, there was an increase in STEMI incidence for every 1 μg/m^3^ increase PM_10_ (Figure S1–5). In subgroup analyses used to evaluate the risk of cardiogenic shock with AP exposure, it is shown that there was a significant association between increasing cardiogenic shock complication rate and for each 1 ppb increase of NO_2_ in patients with no history of PCI, for each 1 μg/m^3^ increase of PM_10_ in patients with prior IHD or without PCI treatment, and for each 1 ppb increase in SO_2_ with HF patients or prior IHD (Figure S6–10).

## Discussion

We performed a large registry-based analysis to evaluate the association between long-term exposure to AP. In this study, using a nationwide prospective clinical registry, we found that increased concentrations of AP increased the risk of STEMI compared with that of NSTEMI and increased the risk of cardiogenic shock complication, which is related to overall mortality. The results of this study showed that long-term exposure to high levels of PM_10_ is associated with an increased risk of STEMI. Moreover, this study demonstrates that PM_10_ and SO_2_ may impact on the development of cardiogenic shock complication in patients with AMI.

Air pollutants comprise complex mixtures that are compounded with gases, including SO_2_, NO_2_, CO, O_3_, and PM, including PM_10_ and PM_2.5_.^15^ Although it may intuitively seem that AP poses a health risk mostly in the form of respiratory disease, many epidemiological and clinical studies have suggested that the majority of the adverse effects of AP are associated with the cardiovascular system.^16–18^ Previous studies demonstrad that AP exposure is associated with endothelial injury and inflammation, indicating that it can trigger cardiovascular events.^19–21^

Regarding the association between AP and CVD, many studies have demonstrated that short-term exposure to AP increases the incidence of an acute coronary syndrome (ACS).^22–24^ In contrast, the present study investigated the effects of long-term exposure to AP, which was the major novelty of our research. The number of studies on the long-term effects of AP exposure are increasing. The ESCAPE (European Study of Cohorts for Air Pollution Effects) study, the increase in PM_10_ and PM_2.5_ during the long-term follow-up period increases the risk of ACS.^25^ However, these studies mainly focus on mortality or overall clinical outcome.^17, 25–27^ Researchers have rarely compared the risk of developing STEMI with that of NSTEMI. To the best of our knowledge, this is the first study to demonstrate a long-term association between AP exposure and the relative incidence of STEMI compared with that of NSTEMI in the Asia-Pacific region.

Our present study was shown that the risk of developing STEMI increased compared to NSTEMI according to the 1-year average PM_10_ concentration before symptom onset. These results indicate that STEMI contributes more than NSTEMI to an increased risk of MI according to the AP concentration. Studies on short-term exposure have reported that elevated AP exposure highly triggers the development of STEMI compared with that of NSTEMI.^28, 29^ However, some studies have also reported greater incidence of NSTEMI than that of STEMI due to AP exposure^30^. The inconsistent results can be attributed to differences in exposure periods, geographic location, pollutant concentration level, study population, and statistical methods used for analysis.^8, 10^

In addition, this study are meaningful in that we clarified the effect of AP exposure on the risk of severe complications of cardiogenic shock. Our main findings showed that an increased AP concentration was associated with an increase incidence of cardiogenic shock complication. Cardiogenic shock occurs in approximately 5–13% of AMI patients.^31^ Moreover, AMI itself was an important etiology contributing to 80% incidences of cardiogenic shock.^32^ Cardiogenic shock is associated with poor prognosis for high rate of adverse events even with appropriate treatment, with an in-hospital mortality of 20–40% and a 1-year mortality rate of up to 50%.^31^

Although the pathophysiology of cardiogenic shock is not fully understood, it is known that the systemic inflammatory response, release of inflammatory cytokines, and increase in the concentration of nitric oxide (NO) are involved in inappropriate vasodilation after peripheral vascular constriction to compensate for the reduction in myocardial contractility.^33, 34^ Exposure to AP is related to oxidative stress and systemic inflammation^14, 16–18^ and it adversely affects vascular homeostasis through the production of superoxide and the uncoupling of NO synthase.^35^ These results add evidence for the development of cardiogenic shock and its subsequent poor prognosis.

In our previously published study, the 1-year average AP concentration before the onset of symptoms was associated with an increase in 30-day short-term mortality.^13^ Studies using the same registry reported that in the STEMI group, there was a higher incidence of short-term mortality and cardiogenic shock compared with the incidence found in the NSTEMI group, which is highly correlated with an increase in the risk of cardiac death.^31, 36^ This study strongly suggests that reducing exposure to high concentrations of AP, not only in the high-risk group but also in the low-risk group of AMI, is necessary to reduce the occurrence of potential MI and mortality, even if STEMI appears to be relatively safe because of its younger age and lower co-morbidity rates than NSTEMI. These efforts should be accompanied by policy strategies and clinical practice.

This study has several limitations. First, because of the limited sampling data available for PM_2.5_, the associations with clinical events may have been relatively low. Exposure data for PM_2.5_, were available for only one year, 2015, the last year of this research period. Evidence suggests that PM size is related to cardiovascular morbidity and mortality.^27, 37^ Further studies with new data are needed to evaluate the impact of PM_2.5_ in AMI patients in the future with our KAMIR data with later than 2015 registry database. Second, because the patients’ addresses were not available, the direct exposure level determined for the patients could be incorrect. It was assumed that patients were admitted to a nearby emergency room at the time of symptom onset. However, some patients who visited other local hospitals or were transferred may have been misclassified, requiring careful interpretation of the results. Third, because of the limitations of the study design, although confounding factors were adjusted for, the results for residential addresses and socioeconomic variables should be carefully considered. We aimed to assess adjusted adequate variables which could be potentially relevant factors using a multivariable model. Finally, although data used in this research were collected by the attending hospitals well-trained, multicenter registry is need to interpret with consideration for several characteristics such as a gap among each hospitals and data such as input errors and misclassification.

In conclusion, we observed that high concentration of air pollutants, particularly of PM_10_, which is an environmental risk factor was associated with an increased incidence of STEMI. Moreover, PM_10_ and SO_2_ levels were risk factors for in-hospital cardiogenic shock complication after MI. This study emphasizes on the need of developing a policy-level strategy and clinical efforts to reduce AP exposure and prevent the incidence of STEMI and severe cardiovascular complications.

## Data Availability

Data cannot be shared publicly because of the KAMIR group policy. Data are available from the Chonnam National University Hospital Institutional Data Access / Ethics Committee (contact via research manager) for researchers who meet the criteria for access to confidential data. Contact information: Research manager: Kyung Hoon Cho, Telephone: +82 62-220-5272

## Acknowledgements

This study was done with the support of Korean Circulation Society (KCS) to commemorate the 50th Anniversary of KCS. The KAMIR study group of the KSC was as follows: Korea University Guro Hospital, Seoul, Korea (Seung-Woon Rha), Gachon University Gil Medical Center, Incheon, Korea (Tae Hoon Ahn), Wonju Severance Christian Hospital, Wonju, Korea (Junghan Yoon), Seoul National University Hospital, Seoul, Korea (Hyo-Soo Kim), Seoul St. Mary’s Hospital, Seoul, Korea (Ki-Bae Seung), Samsung Medical Center, Seoul, Korea (Hyeon-Cheol Gwon), Kyungpook National University Hospital, Daegu, Korea (Shung Chull Chae), Kyung Hee University Hospital at Gangdong, Seoul, Korea (Chong-Jin Kim), Pusan National University Hospital, Busan, Korea (Kwang Soo Cha), Yeungnam University Medical Center, Daegu, Korea (Jung-Hee Lee), Jeonbuk National University Hospital, Jeonju, Korea (Jei Keon Chae), Jeju National University Hospital, Jeju, Korea (Seung-Jae Joo), Seoul National University Bundang Hospital, Seongnam, Korea (Chang-Hwan Yoon), Keimyung University Dongsan Medical Center, Daegu, Korea (Seung-Ho Hur), Chungnam National University Hospital, Daejeon, Korea (In-Whan Seong), Chungbuk National University Hospital, Cheongju, Korea (Kyung-Kuk Hwang), Inje University Haeundae Paik Hospital, Busan, Korea (Doo-Il Kim), Wonkwang University Hospital, Iksan, Korea (Seok Kyu Oh), Gyeongsang National University Hospital, Jinju, Korea (Jin-Yong Hwang), and Chonnam National University Hospital, Gwangju, Korea (Myung Ho Jeong).

## Supplemental Material

Figures S1–S10

## Supplemental data

**Figure S1.**
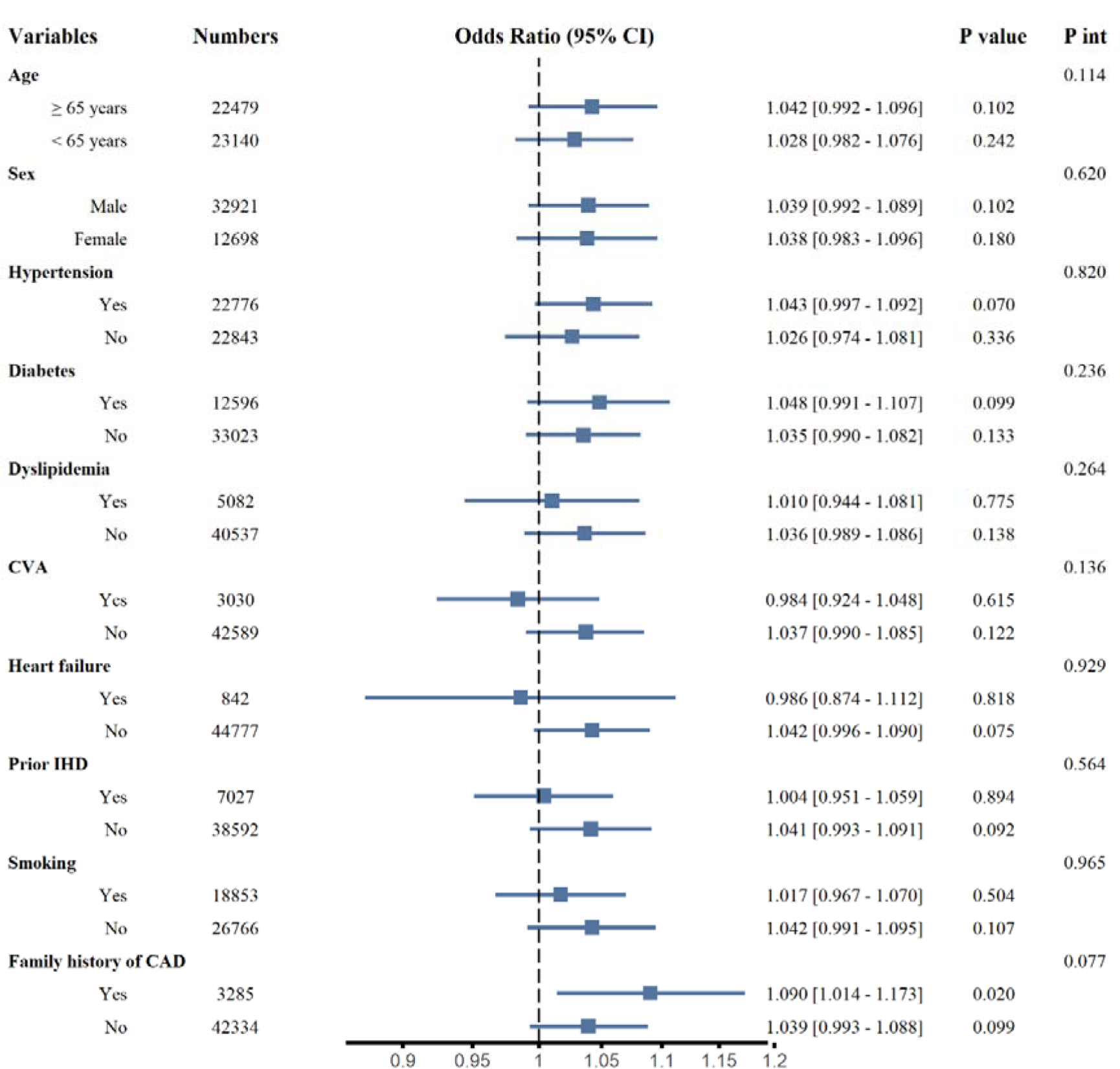
Subgroup analysis for the adjusted odds ratio and 95% confidence interval of the incidence of STEMI compared with NSTEMI according to an increase of 1 part per billion SO_2_ before the onset of symptoms.

**Figure S2.**
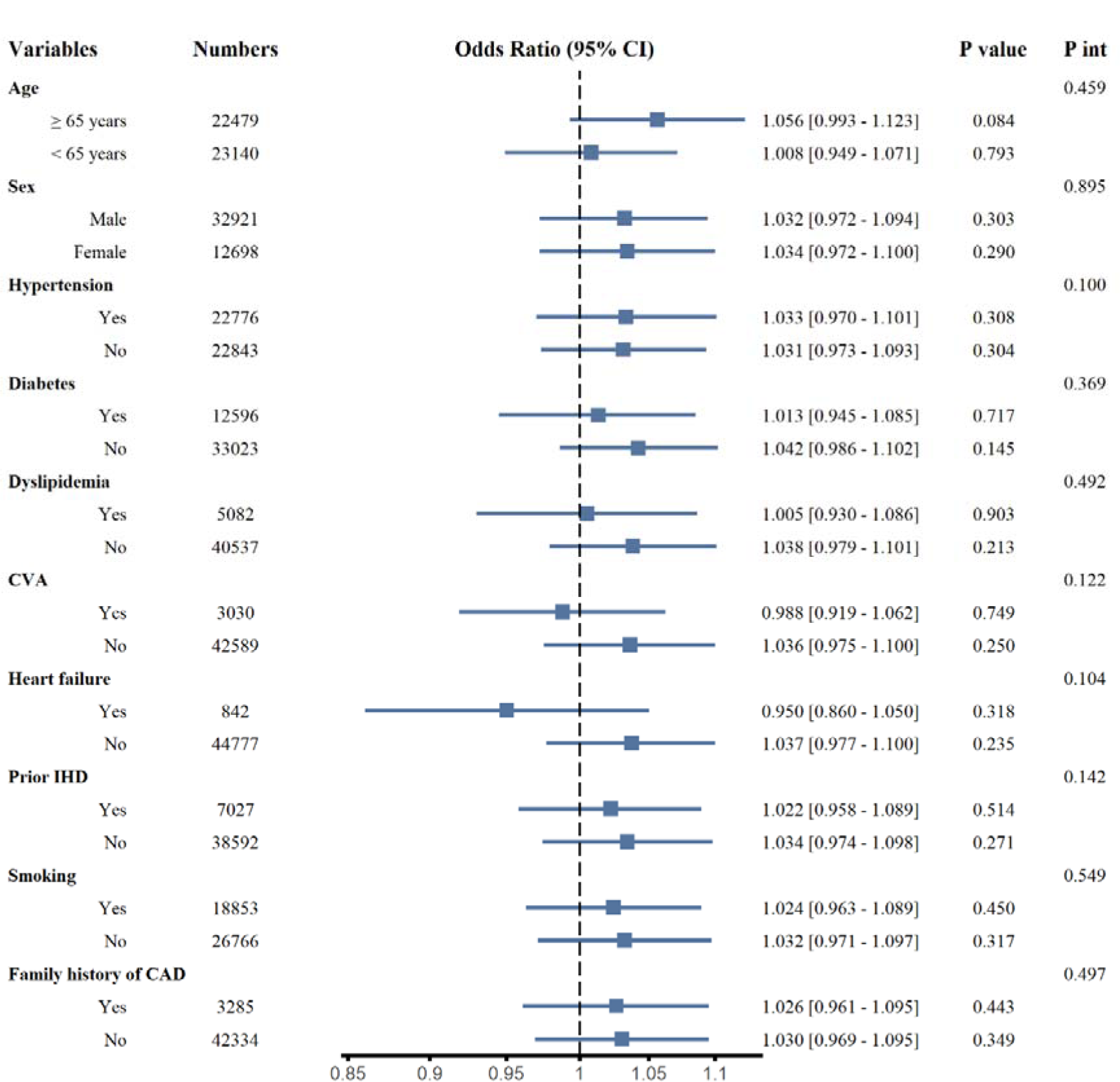
Subgroup analysis for adjusted odds ratio and 95% confidence interval of the incidence of STEMI compared with NSTEMI according to an increase of 0.1 part per million CO

**Figure S3.**
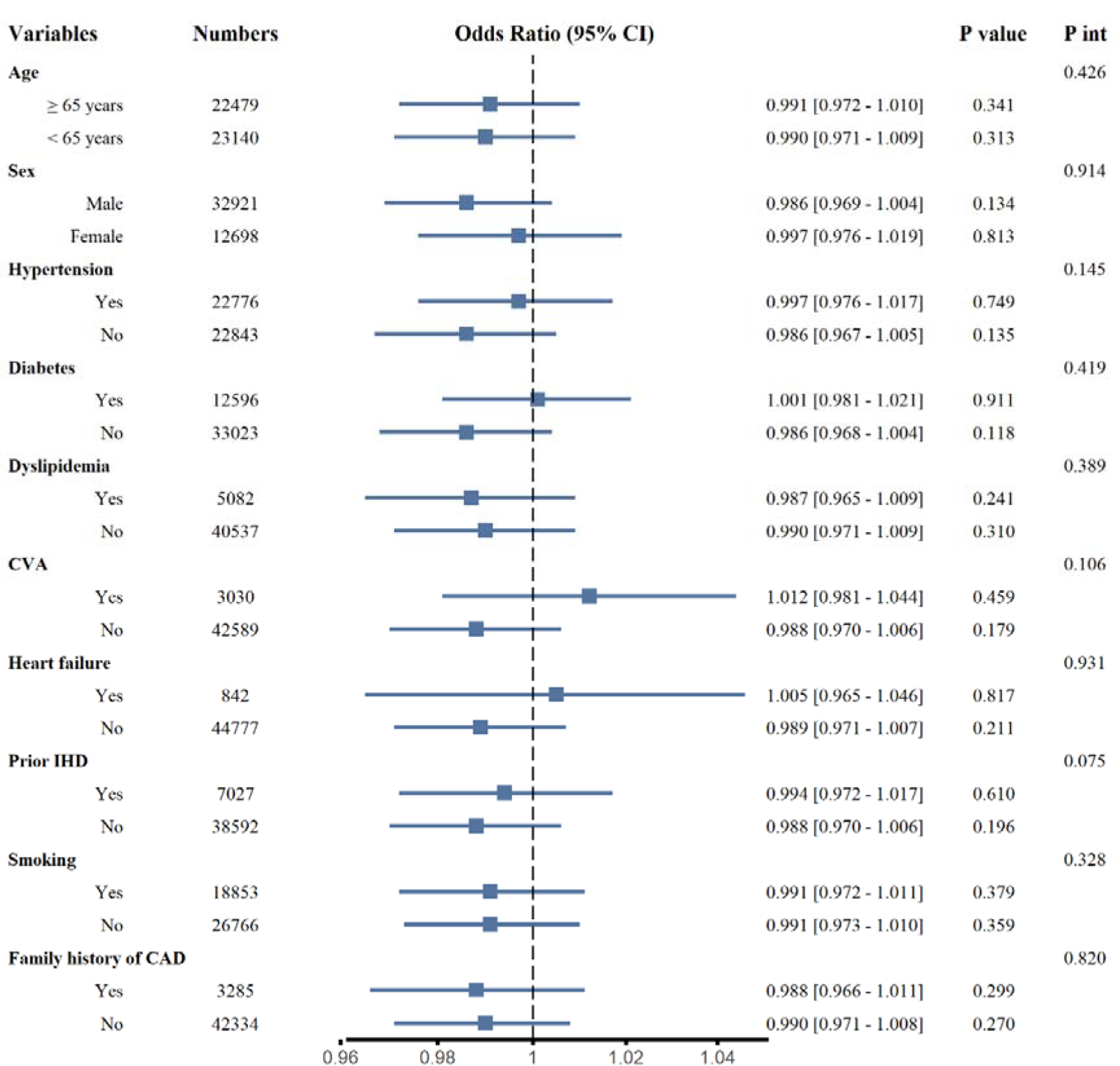
Subgroup analysis for the adjusted odds ratio and 95% confidence interval of the incidence of STEMI compared with NSTEMI according to an increase of 1 part per billion O_3_ before the onset of symptoms.

**Figure S4.**
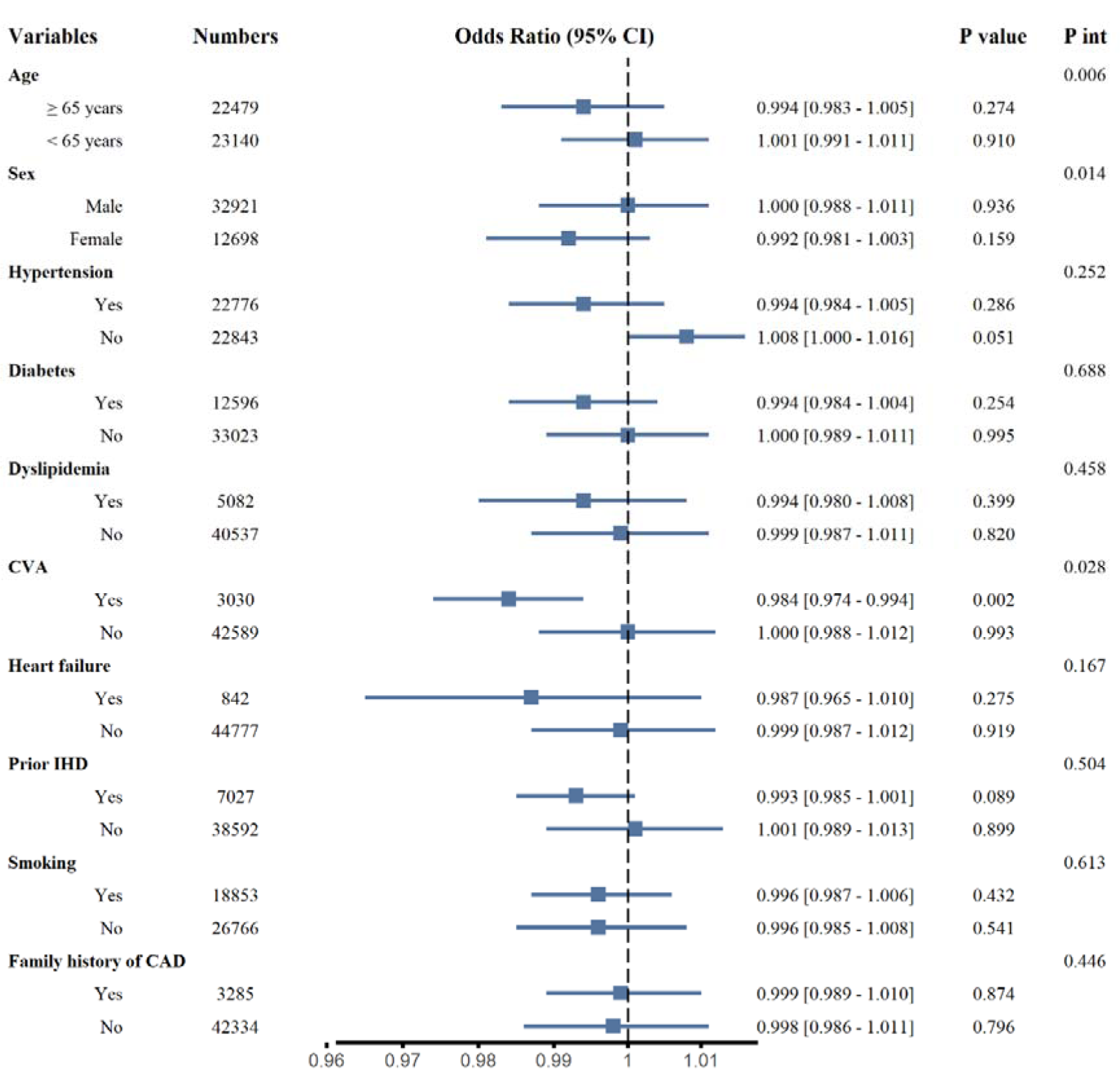
Subgroup analysis for the adjusted odds ratio and 95% confidence interval of the incidence of STEMI compared with NSTEMI according to an increase of 1 part per billion NO_2_ before the onset of symptoms.

**Figure S5.**
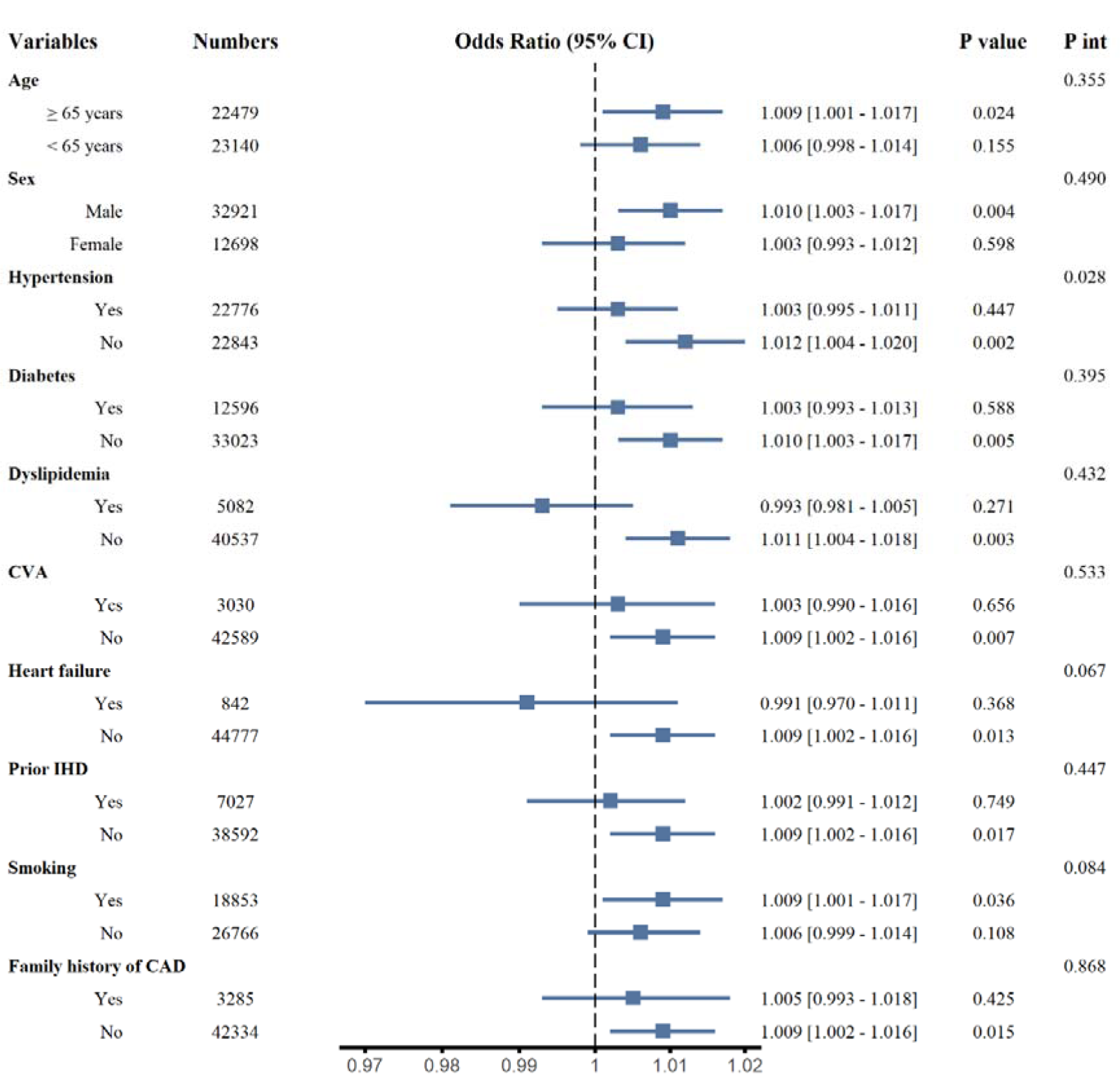
Subgroup analysis for the adjusted odds ratio and 95% confidence interval of the incidence of STEMI compared with NSTEMI according to an increase of 1 µg/m^3^ PM_10_ before the the symptom date.

**Figure S6.**
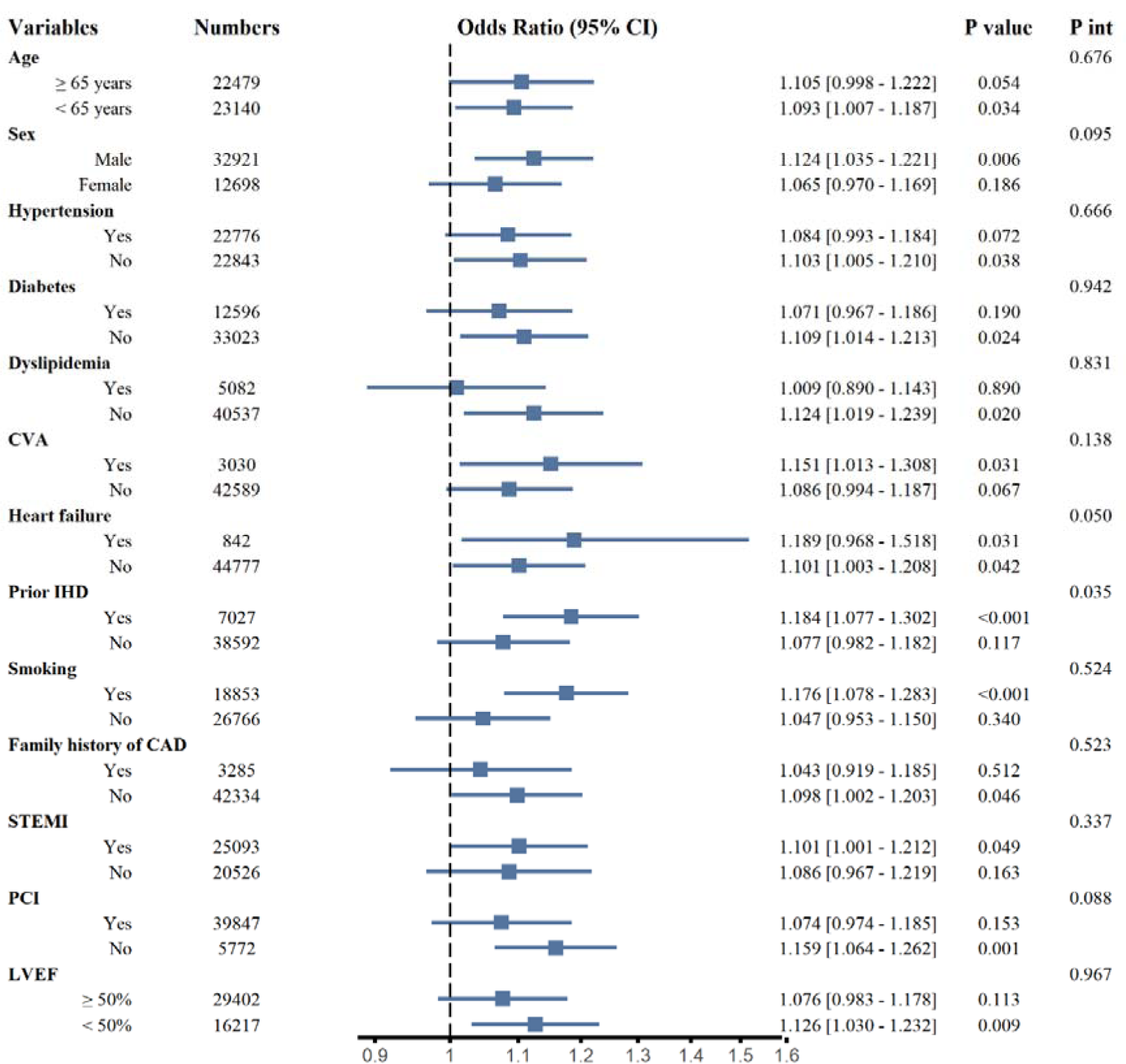
Subgroup analysis for the adjusted odds ratio and 95% confidence interval of the incidence of in-hospital cardiogenic shock according to an increase of 1 part per billion SO_2_ before symptom onset.

**Figure S7.**
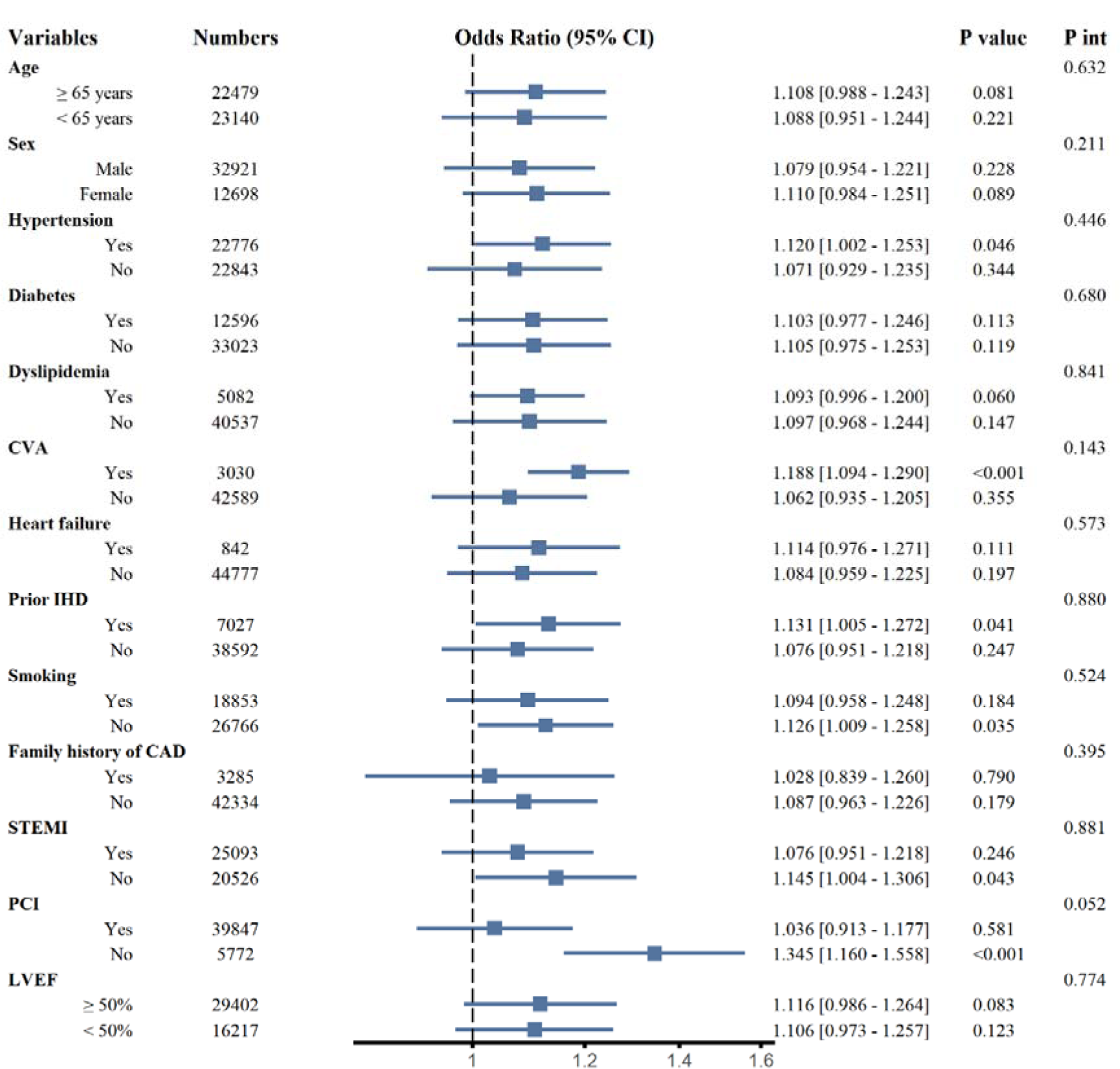
Subgroup Analysis for adjusted odds ratio and 95% confidence interval of the incidence of in-hospital cardiogenic shock according to an increase of 0.1 part per million CO before symptom date.

**Figure S8.**
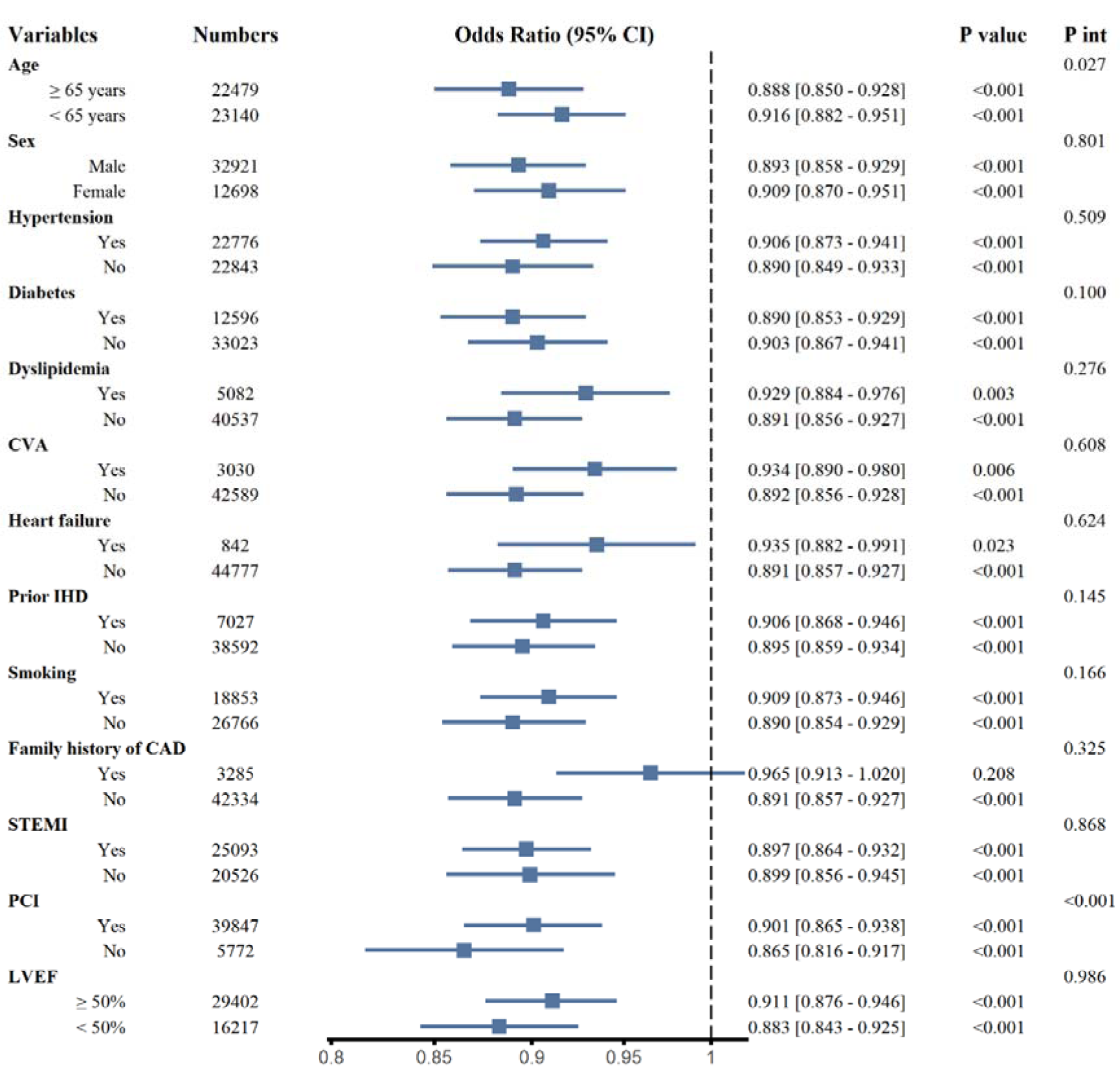
Subgroup analysis for the adjusted odds ratio and 95% confidence interval of the incidence of in-hospital cardiogenic shock according to an increase of 1 part per billion O_3._

**Figure S9.**
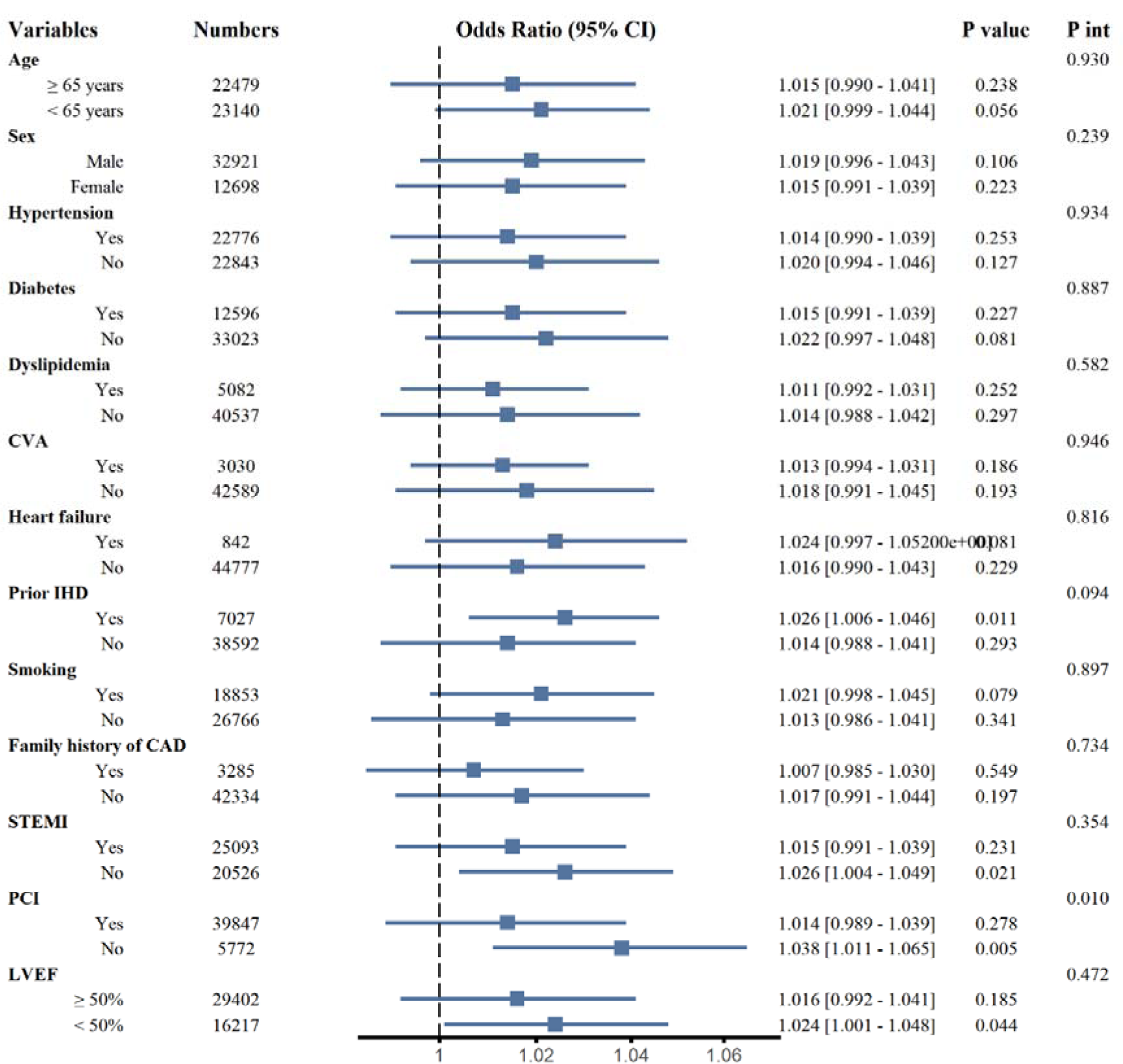
Subgroup analysis for the adjusted odds ratio and 95% confidence interval of the incidence of in-hospital cardiogenic shock according to an increase of 1 part per billion NO_2._

**Figure S10.**
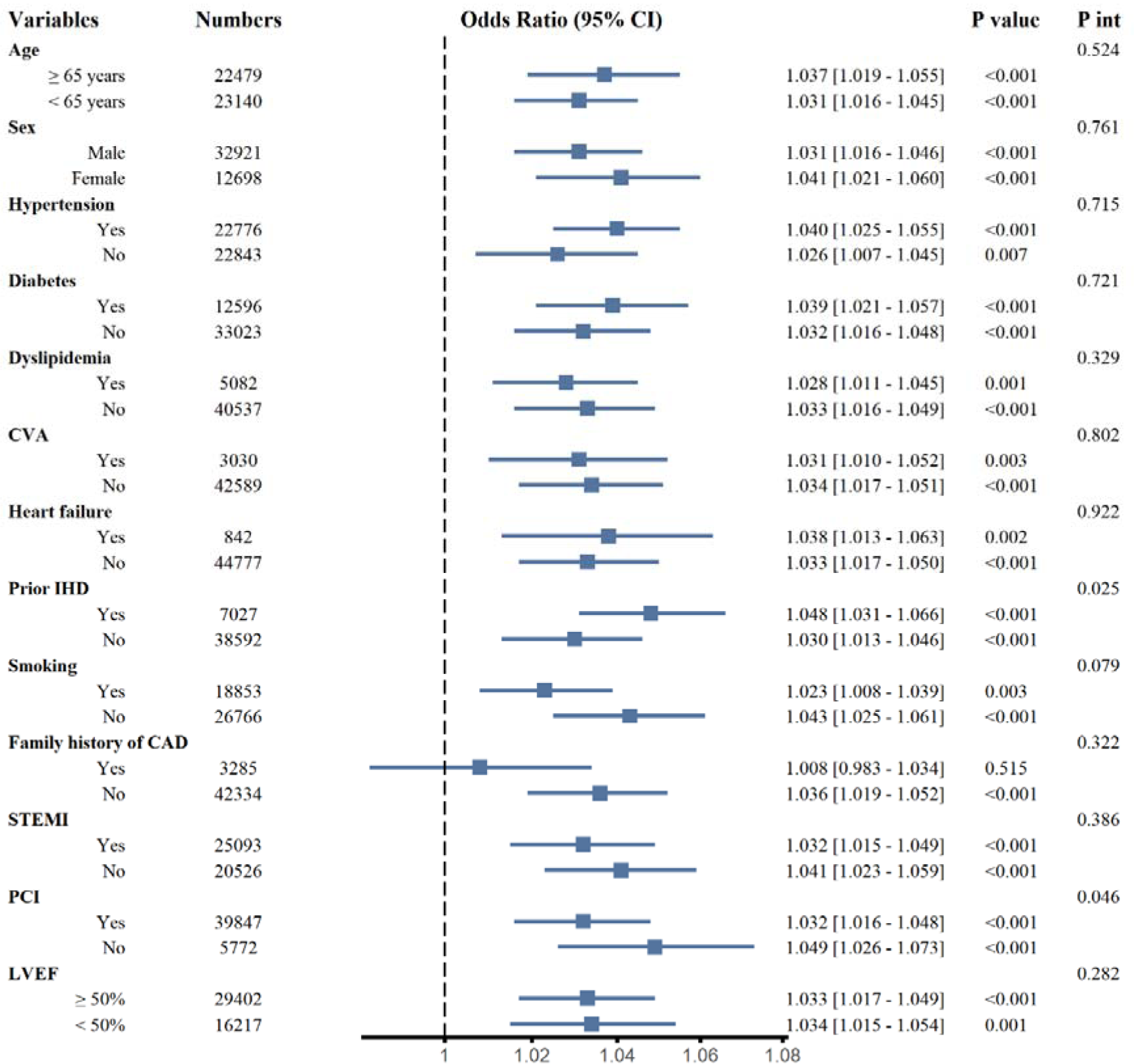
Subgroup analysis for the adjusted odds ratio and 95% confidence interval of the incidence of in-hospital cardiogenic shock according to the increase 1 µg/m^3^ PM_10_ before the symptom date.

